# Regional ventilation characteristics during non-invasive respiratory support in preterm infants

**DOI:** 10.1101/2020.08.04.20168575

**Authors:** J Thomson, CM Rueegger, EJ Perkins, PM Pereira-Fantini, O Farrell, LS Owen, DG Tingay

## Abstract

**Objective:** To determine the regional ventilation characteristics during non-invasive ventilation in stable preterm infants. The secondary aims were to explore the relationship between indicators of ventilation homogeneity and other clinical measures of respiratory status.

**Design:** Prospective observational study.

**Setting:** Two tertiary neonatal intensive care units.

**Patients:** Forty stable preterm infants born <30 weeks gestation receiving either continuous positive applied pressure (n=32) or nasal high-flow cannualae (n=8) at least 24 hours after extubation at time of study.

**Interventions:** Continuous electrical impedance tomography imaging of regional ventilation during 60-minutes of quiet breathing on clinician-determined non-invasive settings.

**Main outcome measures:** Gravity-dependent and right-left centre of ventilation (CoV), percentage of whole lung tidal volume by lung region, and percentage of lung unventilated were determined for 120 artefact-free breaths/infant (4770 breaths included). Oxygen saturation, heart and respiratory rates were also measured.

**Results:** Ventilation was greater in the right lung (mean (SD) 69.1 (14.9)%) total tidal volume and the gravity-nondependent lung; ideal-actual CoV 1.4 (4.5)%. The central third of the lung received the most tidal volume, followed by the non-dependent and dependent regions (p<0.0001 repeated measure ANOVA). Ventilation inhomogeneity was associated with worse SpO_2_/FiO_2_, (p=0.031, r^2^ 0.12; linear regression). In those infants that later developed bronchopulmonary dysplasia (n=25) SpO_2_/FiO_2_ was worse and non-dependent ventilation inhomogeneity greater than in those that did not (both p<0.05; t test Welch correction).

**Conclusions:** There is high breath-by-breath variability in regional ventilation patterns during NIV in preterm infants. Ventilation favoured the gravity-nondependent lung, with ventilation inhomogeneity associated with worse oxygenation.

## INTRODUCTION

Clinical guidelines now recommend non-invasive ventilation (NIV) as initial respiratory support for preterm infants with respiratory failure.[1] NIV is simpler to use than support via an endotracheal tube, well tolerated and confers short and long-term lung protective benefits.[2] In a meta-analysis of clinical trials, the use of NIV resulted in a small reduction in bronchopulmonary dysplasia (BPD).[2] Unfortunately, population follow-up studies have not demonstrated a reduction in BPD, with poorer late-childhood respiratory outcomes reported in recent cohorts, compared with earlier cohorts where NIV was less commonly used.[3-5] Beyond the delivery room, NIV is usually provided with either continuous positive airway pressure (CPAP) or high-flow nasal cannualae (nHF). The indications for use, and specific CPAP and nHF strategies vary widely,[1,6] potentially explaining some of the discrepancy between trial and clinical respiratory outcomes.

BPD is a multi-factorial progressive process that results from injurious respiratory support delivered to an immature lung with reduced alveolar capacity.[7] Injurious ventilation states can be caused by inadequate lung volume (atelectasis) or excessive tidal ventilation (volutrauma), both of which can generate damaging mechanotransductive forces resulting in increased oxygen needs and a biotrauma response.[8-10] Understanding where these events are occurring in the lung requires methods of monitoring volume state, ideally on a breath-by-breath basis.[10,11] However, continuous monitoring during NIV is limited to peripheral measures of gas exchange and clinical observation of work of breathing.

Studies in adults and preterm animals have consistently shown that ventilation is rarely uniform within the diseased lung.[10,12,13] In preclinical studies heterogeneous (non-uniform) ventilation worsens oxygenation and creates complex injury events, potentially allowing different volume states to occur in different lung regions simultaneously.[9,10,13] In animal studies, improving ventilation uniformity reduces acute markers of lung injury.[10,13-15] Currently, clinicians have no bedside method of assessing ventilation uniformity in preterm infants, limiting the ability to determine whether NIV settings are lung protective or injurious. This hampers the ability of respiratory treatments to be modified before the cascade of events causing BPD develops.[4]

Recently, electrical impedance tomography (EIT) systems for infants have become available.[11,16] EIT is a method of breath-by-breath imaging of regional ventilation homogeneity and relative tidal volume (V_T_) that is ideally suited to preterm infants, being non-invasive, radiation-free and continuous during all forms of support.[11,17] Limited EIT data exists in preterm infants receiving NIV,[11,18,19] especially with regards to associations with clinical status. The aim of this study was to determine the regional ventilation characteristics during non-invasive ventilation in stable preterm infants. The secondary aims were to explore the relationship between indicators of ventilation homogeneity and other clinical measures of respiratory status, including BPD. This is the first step in understanding the potential clinical utility of measures of ventilation homogeneity to guide lung-protective management in the NICU.

## METHODS

This prospective observational study was performed in the Neonatal Intensive Care Units (NICUs) of the Royal Children’s Hospital and Royal Women’s Hospital, Melbourne, Australia between November 2016 to August 2017. Prospective written informed parental consent was provided.

Infants receiving either CPAP or nHF were eligible if they were born <30 weeks of gestation, were <36 weeks corrected gestation at time of study, and had been extubated for at least 24 hours. Infants were not studied if they had at least one period of >4 hours without any respiratory support, NIV was being used for a primary cardiac disease or to support intrinsic or iatrogenic suppressed central respiratory drive on day of study, had fragile skin or were deemed too medically unstable to handle. The mode and settings of NIV and infant body position were determined by the clinician and were not altered during study.

An ultrasound gel-coated, non-adhesive, 32-electrode neonatal EIT belt (SenTec AG, Landquart, Switzerland) was placed around the infant’s chest at nipple level during a period of clinical handling using our previously described methods.[16,20-23] The belt was connected to the SenTec Pioneer EIT system and electrode signal quality confirmed. After allowing the infant to settle, four 10-minute EIT recordings were performed at 48 frames/s during 60-minutes of quiet breathing. Peripheral capillary oxygen saturation (SpO_2_), heart rate (HR) and respiratory rate (RR) were measured continuously (Intellivue MD70 monitor, Phillips Healthcare, Eindhoven, Netherlands) and values recorded minutely.

Raw EIT data were reconstructed in accordance with standardised guidelines[11,24] with an anatomically correct custom-made algorithm of the human chest to exclude impedance change outside the right and left lung boundaries using the manufacturer’s software package (ibeX, SenTec AG). Once reconstructed, each of the 10-minute recordings were manually reviewed using a standardised approach to identify the first 30 consecutive artefact-free inflations (total 120 inflations/infant) to include for analysis.[17,19,22] For each selected inflation functional EIT images of the distribution of V_T_ were created, and data extracted from the whole lung, right and left lung, and three gravity-dependent regions of interest (ROIs); gravity-nondependent (ND), central (C) and gravity-dependent (D).[11] The following measures of ventilation homogeneity were generated for each inflation: 1) the relative V_T_ within each ROI as a percentage of total V_T_ in the lung and as a ratio of anatomical size of each ROI (%V_T_ in the gravity-nondependent, central and gravity-dependent ROI only analysed for prone and supine infants; Figure 1C); 2) the centre of ventilation (geometric mean of V_T_ distribution along a single plane) along the right-left (CoV_RL_), and gravity-nondependent to dependent plane,[11,25] and 3) the percentage of unventilated lung regions.[11] To adjust for various body positions the gravity-dependent CoV was referenced to the CoV value representing homogenous (ideal) ventilation for that body position (calculated by ibeX),[11] generating the ideal CoV to actual CoV difference (CoV_I-A_). CoV_I-A_ 0 = homogenous ventilation; >0 ventilation inhomogeneity favouring the gravity-nondependent lung; <0 ventilation inhomogeneity favouring the dependent lung (Online Supplementary Material [OSM] Figure 1).

**Figure 1.**
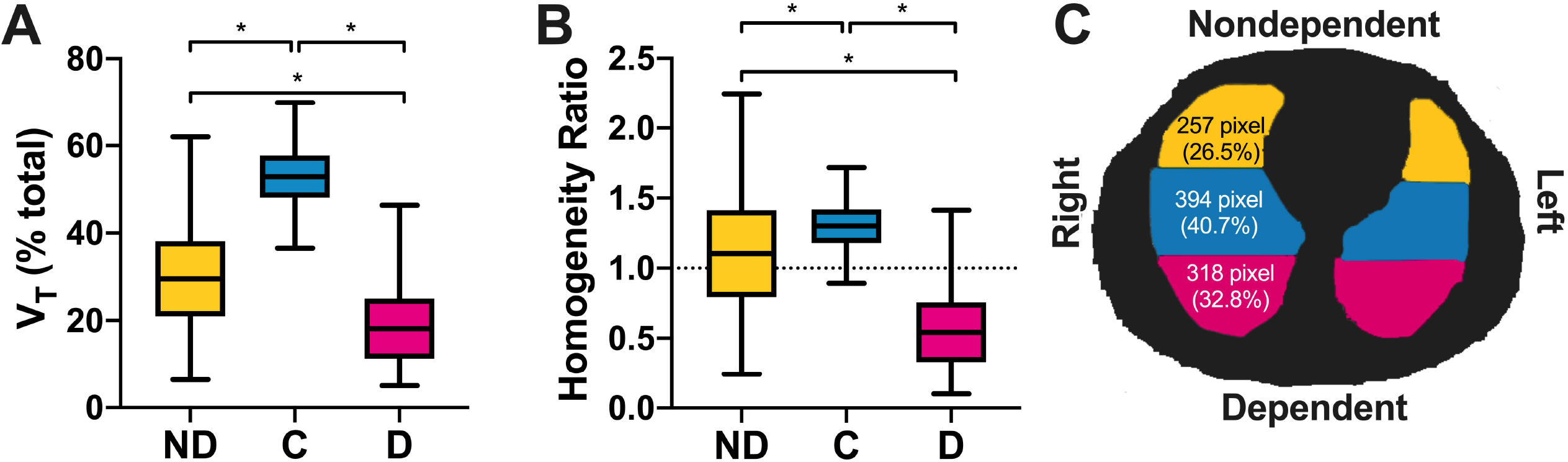
**A**. Proportion (%) of V_T_ occurring in the gravity-nondependent (ND; yellow), central (C; blue) and gravity-dependent (D; pink) ROI thirds of the lungs. **B**. Proportion of V_T_ occurring in the thirds of the chest (using same symbols as Panel A) expressed as a ratio of the anatomical contribution (ventilation homogeneity ratio) of the gravity-nondependent (26.5%), central (40.7%) and gravity-dependent (32.8%) ROI thirds to the entire lung regions imaged. This ratio accounts for the different anatomical size of each region, and the different anatomical regions representing gravity-dependent and nondependent when prone and supine. A value >1 indicates that V_T_ was relatively greater in that region compared to the size of the region, <1 V_T_ relatively less than the size of the region, and 0 = uniformity. **C**. Schematic of the shape of the lungs within the field of EIT imaging demonstrating the gravity-nondependent, central and gravity-dependent regions and the absolute number of pixels (and percentage of total lung regions) that each region contains. Values represent the combination of the right and left lung for each ROI. Infants only included if in supine or prone position, and adjusted for position (n=34). All boxes SD and line mean, error bars range. * p<0.0001 Tukey post test (repeated-measure ANOVA)

The true pattern of ventilation in the preterm lung is unknown. Based on previous EIT studies of preterm infants, 18 infants were required to detect a clinically meaningful 2% difference in gravity-dependent CoV (assuming 3% SD, power 80% and alpha error 0.05).[17] A convenience sample size of 40 infants was chosen to allow exploration of pre-allocated variables that may impact ventilation pattern; type of NIV, oxygen deficit at study (measure of acute disease severity), and indicators of developmental lung state (gestational age (GA) at birth, corrected GA, age at time of study). Finally, all these variables were compared with subsequent bronchopulmonary dysplasia (BPD) status at 36-weeks corrected gestation (modified Walsh criteria).[26] The relationship between these variables and CoV measures were compared with linear regression. All data were tested for normality, and additional analysis performed with appropriate categorical or continuous test. Welch correction or robust error cluster analysis (by infant) were applied as required. Statistical analysis was performed using Prism (v7.0, GraphPad, CA, USA) and a p value <0.05 considered significant.

## RESULTS

During the study period, 109 infants were eligible. Forty were studied and provided complete data sets (4770 breaths; OSM Figure 2). The characteristics of the infants studied are described in Table 1. At time of study 32 infants were receiving CPAP, and 8 nHF. The range of CPAP distending pressure was 5-10 (median 7) cmH_2_O, and nHF flow rate 4-6 L/min. Infants receiving CPAP were more immature at birth, more likely to have been intubated and received exogenous surfactant, and had greater oxygen deficit and respiratory rate at time of study. Thirty-one infants were studied prone (30 CPAP), three supine (0 CPAP) and six were lateral (2 CPAP). The mean (SD) inspiratory time was 0.40 (0.14) s, with a mean (95% CI) difference between CPAP and nHF of 0.07 (−0.01, 0.15) s (t-test).

**Table 1.** Infant characteristics.

|  | **All; n=40** | **CPAP; n=32** | **nHF; n=8** |
| --- | --- | --- | --- |
| **Demographic Characteristics** |  |  |  |
| GA at birth (completed weeks) | 27 (2) | 26 (2) | 28 (2)* |
| Corrected GA (completed weeks) | 31 (2) | 31 (2) | 31 (1) |
| Postnatal age (days) | 28 (6, 67) | 33 (8, 67) | 12 (6, 66) |
| Birth weight (g) | 947 (226) | 903 (178) | 1123 (317) |
| Weight at study (g) | 1332 (311) | 1336 (339) | 1316 (168) |
| Female, n (%) | 19 (48%) | 15 (47%) | 4 (50%) |
| Twins, n (%) | 10 (25%) | 8 (25%) | 2 (20%) |
| **Respiratory Characteristics** |  |  |  |
| Intubated prior to study, n (%) | 35 (88%) | 30 (94%) | 5 (63%)<sup>†</sup> |
| Exogenous surfactant therapy, n (%) | 34 (85%) | 30 (94%)<sup>#</sup> | 4 (50%)<sup>††</sup> |
| Caffeine on day of study, n (%) | 37 (93%) | 29 (91%) | 8 (100%) |
| Bronchopulmonary Dysplasia, n (%) | 25 (63%) | 22 (69%) | 3 (38%) |
| **Cardiorespiratory status at study commencement** |  |  |  |
| FiO<sub>2</sub> | 0.30 (0.21, 0.45) | 0.30 (0.21, 0.45) | 0.21 (0.21, 0.28)<sup>‡</sup> |
| SpO<sub>2</sub> (%) | 93 (4) | 93 (3) | 96 (5) |
| SpO<sub>2</sub>/FiO<sub>2</sub> | 345 (83) | 321 (69) | 443 (55)** |
| Heart rate (bpm) | 163 (12) | 163 (13) | 165 (10) |
| Respiratory rate (breaths per min) | 65 (30, 92) | 68 (30, 92) | 51 (37, 70) |

All data mean (SD), median (range) or number (%).
*Abbreviations:* GA, gestational age; FiO<sub>2</sub>, Fraction of inspired Oxygen; SpO<sub>2</sub>, Peripheral oxygen saturation; bpm, beats per minute.

**Figure 2.**
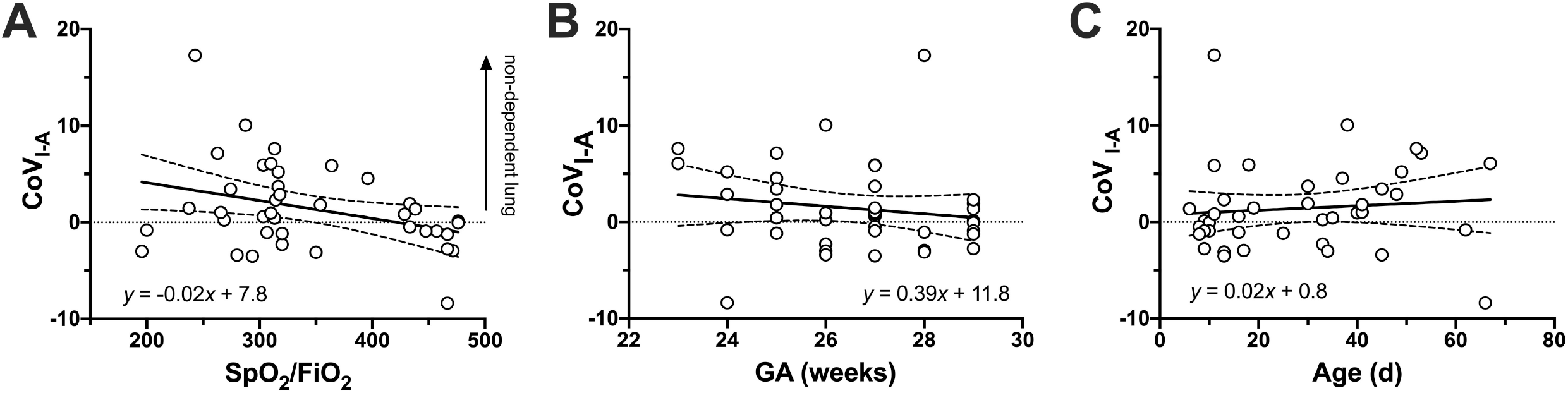
Relationship between the gravity-dependent uniformity of ventilation (CoV_I-A_) SpO_2_/FiO_2_ at start of study (**A**; p=0.031, R^2^=0.12, linear regression), gestational age (GA, **B**; p=0.33, R^2^=0.02) and age at study (**C**; p=0.56, R^2^=0.01) for all 40 infants. A CoV_I-A_ of 0 (dashed line) represents uniform ventilation, and a value >0 relatively greater V_T_ in the gravity-nondependent lung. Greater SpO_2_/FiO_2_ values represents better oxygenation. Solid line represents line of best fit (equation in panel; linear regression) and dashed lines the 95% CI.

### Distribution of ventilation

Table 2 describes the CoV data. Ventilation was greater in the right lung, with a mean (SD) 69.1 (14.9)% of total lung V_T_ during each inflation occurring in the right lung. There was no difference between CPAP and nHF; 3.8 (−3.1, 10.7)% of V_T_.

**Table 2.** Uniformity of ventilation.

|  | **All (n=40)** | **CPAP (n=32)** | **nHF (n=8)** | **Difference** |
| --- | --- | --- | --- | --- |
| **CoV<sub>RL</sub> (%)** | 41.2 (7.4)% | 40.7 (8.1)% | 43.2 (3.5)% | 2.5 (-1.4, 6.4)% |
| *Ideal (uniform) ventilation occurs at 46% of the distance from the most right-hand (0%) to left-hand (100%) lung regions.* |  |  |  |  |
| **CoV<sub>I-A</sub>** | 1.4 (4.5) | 2.0 (4.5) | -1.1 (3.6) | 3.1 (-0.1, 6.3) |
| *Ideal (uniform) ventilation occurs at 0, values >0 indicate relatively greater V<sub>T</sub> in the gravity-nondependent lung, and <0 the dependent lung* |  |  |  |  |
| **Percentage of lung regions without any apparent tidal ventilation** |  |  |  |  |
| **Whole lung** | 12.5 (5.7)% | 12.3 (5.9)% | 13.4 (4.9)% | 1.1 (-3.3, 5.5)% |
| **Non-dependent lung** | 6.9 (3.9)% | 7.1 (4.2)% | 6.2 (2.1)% | 0.8 (-1.3, 3.0)% |
| **Dependent lung** | 5.6 (3.7)% | 5.2 (3.7)% | 7.1 (3.8)% | 2.0 (-1.3, 5.3)% |

All data mean (SD) and differences mean (95% CI); t test with robust error. n = number of infants (4770 breaths total).
*Abbreviations:* CoV<sub>RL</sub>, Centre of ventilation along the right to left plane; CoV<sub>I-A</sub>, ratio of actual to ideal centre of ventilation along the gravity-dependent plane adjusted for body position at time of study.

Overall, the relative V_T_ delivered to the gravity-nondependent, central and gravity-dependent ROI of the chest differed (Figure 1; p<0.0001, repeated-measure ANOVA), even when adjusted for the different anatomical sizes of each ROI (p<0.0001). Ventilation was relatively greater in the non-dependent lung in infants receiving CPAP, and favoured the dependent lung in the eight infants receiving nHF, but the difference was not significant (p=0.058). CoV_I-A_ by each nursing position are shown in OSM Table 1.

#### Relationship between uniformity of ventilation, oxygenation and developmental lung state

There was no relationship between CoV_RL_ and oxygenation (SpO_2_/FiO_2_ ratio), GA at birth and age at study (data not shown). Inhomogeneity of ventilation favouring the non-dependent lung was associated with worse oxygenation (Figure 2A). There was no relationship between CoV_I-A_ and GA, age at study, corrected GA, weight, birth weight and PEEP (Figure 2B and C, OSM Table 2).

#### BPD status

Twenty-five infants (62.5%) were assessed as later having BPD. Infants who met the criteria for BPD were more immature and smaller at birth, but were older, heavier and had worse oxygenation at time of EIT study (Table 3). At the time of study, ventilation was more heterogenous, being greater in the non-dependent lung, and expressing greater variability in those infants who later developed BPD compared with those who did not.

**Table 3.** Outcome measures at time of study and later bronchopulmonary dysplasia status.

|  | **No BPD (n=15)** | **BPD (n=25)** | **Difference** |
| --- | --- | --- | --- |
| **CoV<sub>I-A</sub>** | -0.5 (2.2) | 2.6 (5.1) | 3.1 (0.7, 5.4) |
| **SpO<sub>2</sub>/FiO<sub>2</sub> ratio** | 402 (73) | 311 (69) | 92 (44, 139) |
| **GA at birth (completed weeks)** | 28 (1) | 26 (2) | 2 (1, 3) |
| **Age at study (days)** | 11 (6, 33) | 38 (10, 67) | p<0.0001* |
| **Corrected GA (completed weeks)** | 31 (1) | 30 (1) | 1.5 (0.6, 2.3) |
| **Birthweight (g)** | 1084 (212) | 865 (191) | 219 (79, 360) |
| **Weight at study (g)** | 1193 (253) | 1415 (317) | 221 (36, 406) |
| **PEEP (cmH<sub>2</sub>O)<sup>†</sup>** | 6.8 (1.5) | 7.1 (1.2) | 0.3 (-1.5, 0.8) |

*All data mean (SD) except median (range). All differences mean (95% CI) t test with Welch's*
*correction, except \* Mann-Whitney test. <sup>†</sup> CPAP group only (10 no BPD, 22 BPD).*

## DISCUSSION

Non-invasive ventilation is the most commonly used type of respiratory support for preterm infants, but lacks reliable methods of measuring the impact on lung function. In our observational study we demonstrated that readily available EIT measures of ventilation homogeneity can be used during NIV, and were related to disease and developmental state. Importantly, preterm infants exhibited ventilation inhomogeneity during NIV, with ventilation favouring the gravity-nondependent lung regions, a pattern acknowledged to be injurious in other patient groups.[10,27,28] There was an association between worse oxygenation and relatively greater non-dependent ventilation inhomogeneity, suggesting that EIT may have merit in monitoring respiratory status and guiding care in the NICU.

The primary purpose of this study was to describe the regional ventilation patterns in a representative group of stable preterm infants receiving NIV. We found that ventilation was greater in the right lung and the non-dependent lung, with high breath-by-breath variability. This suggests that ventilation inhomogeneity favouring the non-dependent lung, a well-described characteristic of acute parenchymal lung disease,[19,29-33] persists beyond the period of initial surfactant-deficient RDS, and differs from the predominantly dependent ventilation pattern described in spontaneously breathing term infants.[23,34] Ventilation inhomogeneity creates the potential for opposing mechanisms of ventilator-induced lung injury to occur simultaneously; atelectasis in poorly ventilated regions and volutrauma in over-ventilated, even if the delivered overall V_T_ is appropriate for gas exchange.[9,10,28] Preterm infants often receive NIV for prolonged periods,[5] and ventilation homogeneity is rarely a treatment goal. Our study suggests the risk of injury to the under-developed preterm lung goes well beyond the period of acute RDS, and may have a role in the increasing rates of BPD.[3,5]

As expected, the study population did not have high oxygen needs. Thus, the finding that ventilation distribution along the gravity-dependent plane was associated with oxygenation, but not other parameters, is interesting. Oxygenation is the most commonly used parameter to guide respiratory support in the NICU,[1] but is influenced by non-pulmonary factors, and rarely discriminates beyond identifying treatment failure or suitability of ceasing NIV. EIT measures of ventilation and aeration have been shown to be more accurate at differentiating suitability of pressure settings during high-frequency ventilation and NIV.[18,19,30,35] Recently, EIT-guided PEEP titration, based on achieving ventilation homogeneity, has been demonstrated to improve oxygenation compared to existing approaches in acute respiratory distress syndrome.[33] EIT may have an important role in optimising NIV, especially as setting PEEP/flow levels during NIV in preterm infants still lacks evidence-based approaches.[1] Although the goodness of fit was low (r^2^<0.5), there is a biological plausibility between increased ventilation heterogeneity and worse oxygenation that supports the use of EIT in the NICU and should be validated in larger studies.

The majority of infants were receiving CPAP at the time of study, but 20% were being managed with nHF. Although CoV_RL_ and COV_I-A_ were more homogeneous in the infants studied during nHF these data likely reflect the lack of matching based on type of NIV, and the small number receiving nHF. Consistent with clinical practice in Australia, infants receiving nHF had less severe lung disease before and at the time of study.[5] CPAP and nHF deliver NIV via different mechanisms, but EIT ventilation patterns were the same in a cross-over study of nHF and bubble CPAP.[36] We postulate that the role of spontaneous breathing and disease state will be more important to regional ventilation than type of NIV if applied appropriately.[22]

To date EIT studies in infants have been observational.[11] In part this is due to the need to first describe the respiratory patterns of this poorly understood population in which diseases, lung development, interventions and clinical settings vary.[4,7] Clinicians will require evidence that EIT offers the potential to modulate outcomes before widespread use,[11] as has been demonstrated in animal studies.[10,31,37] Current neonatal EIT systems now have real-time processing and simple non-adhesive belts that can be applied during multiple scenarios, and for prolonged periods.[16,38] EIT has already been proposed as a simple radiation-free method of determining endotracheal tube location and air-leaks.[11,39,40] The observation that gravity-dependent V_T_ inhomogeneity differed between infants that developed BPD and those that did not is intriguing. This is especially so as inclusion in our study was based on feasibility, and a single measurement made at a non-standardised time point. An observation between ventilation inhomogeneity and later BPD status is purely exploratory but provides a justification to further investigate the ability of repeated EIT-measures of ventilation inhomogeneity from birth to predict BPD. The lack of an early biomarker of BPD risk when disease-modifying therapies may still be effective represents the largest single hurdle to meaningful impact on the long-term respiratory consequences of preterm birth.[4,7]

This study has limitations not already highlighted. The eligibility criteria were broad, sample size based on feasibility and sub-group analysis intentionally exploratory. The clinical status of infants was not mandated, and a diverse population was studied. This limits interpretation but, as this is the largest study of NIV ventilation patterns in preterm infants, does not diminish the key finding that highly variable ventilation inhomogeneity is a feature of NIV. Our study will inform the design of larger studies that account for the multi-factorial processes influencing BPD.[7] We used parameters routinely available in current EIT systems. Other measures of heterogeneity, such as the global inhomogeneity index,[11] warrant investigation in preterm infants. We did not standardise infant position during the study, but did adapt our analysis to account for it. The majority of infants were studied prone, limiting the interpretation of the differences in COV_I-A_ seen in the small numbers nursed in other positions. If nursing position does alter ventilation patterns this is important and requires further investigation.

In conclusion, preterm infants generate high breath-by-breath variability in regional ventilation during NIV even when clinically stable. Ventilation favoured the gravity-nondependent lung, with ventilation inhomogeneity associated with worse oxygenation. The degree of ventilation inhomogeneity increases lung injury potential in other populations, and may have use in guiding care and identifying infants at greatest risk of BPD whilst disease risk is modifiable.

### What is already known on this topic

1. Non-invasive ventilation (NIV) is the most commonly used type of respiratory support for preterm infants and offers lung protective benefits in clinical trials.
2. The application of NIV is not standardised. In a recent long-term respiratory follow up study, outcomes had worsened in the era of greater NIV use.
3. Respiratory support that creates heterogenous ventilation within the lungs is known to worsen oxygenation, and may increase lung injury, in adults and preterm animals.

### What this study adds

1. Breath-by-breath tidal ventilation was highly variable within the lungs of stable preterm infants receiving NIV beyond the initial acute disease phase.
2. Ventilation was heterogenous, favouring the right and gravity-nondependent lung. Increased ventilation inhomogeneity was associated with worse oxygenation and a later diagnosis of bronchopulmonary dysplasia.
3. Bedside assessment of ventilation patterns within the lung is possible using modern electrical impedance tomography systems, and offers potential to guide disease-modifying respiratory support.

## Data Availability

Deidentified individual participant data, study protocols and statistical analysis codes are available from three months to 23 years following article publication to researchers who provide a methodologically sound proposal, with approval by an independent review committee (learned intermediary). Proposals should be directed to to gain access. Data requestors will need to sign a data access or material transfer agreement approved by MCRI.

## Competing interests

There are no competing interests to declare.

## Ethics approval

This study was approved by the Royal Children’s Hospital Human Research Ethics Committee (HREC 36159A) and the Royal Women’s Hospital Human Research Ethics Committee (HREC 16-33), in accordance with the National Health and Medical Research Council Statement of Ethical Conduct in Human Research (2007).

## Financial Support

This study is supported by the Victorian Government Operational Infrastructure Support Program (Melbourne, Australia). DGT is supported by a National Health and Medical Research Council Clinical Career Development Fellowship (Grant ID 1053889). CMR is supported by a Swiss National Science Foundation Early Postdoctoral Mobility fellowship (P2ZHP3_161749) and the Milupa Fellowship Award of the Swiss Society of Neonatology. SenTec AG (Landquart, Switzerland) manufactured custom-built EIT belts for infants. All EIT hardware was purchased by MCRI without any restrictions.

## Author contributions

DGT and LSO developed the concept and experimental design. JT, CMR, EJP and DGT were involved in data acquisition and JT, OF and CMR analysed the data. DGT, PP-F, LSO and EP supervised the study. All authors participated in data interpretation. JT and DGT drafted the first manuscript and all authors contributed to editing.

## Data sharing

Deidentified individual participant data, study protocols and statistical analysis codes are available from three months to 23 years following article publication to researchers who provide a methodologically sound proposal, with approval by an independent review committee (“learned intermediary”). Proposals should be directed to to gain access. Data requestors will need to sign a data access or material transfer agreement approved by MCRI.

## Notes

### Competing Interest Statement

The authors have declared no competing interest.

### Clinical Trial

Australian and New Zealand Clinical Trials Registry (ACTRN12616001516471)

### Author Declarations

This study was approved by the Royal Childrens Hospital Human Research Ethics Committee (HREC 36159A) and the Royal Womens Hospital Human Research Ethics Committee (HREC 16-33), in accordance with the National Health and Medical Research Council Statement of Ethical Conduct in Human Research (2007).

### Summary of Updates

Revision following Reviewer comments (ADC FN)

## References

1. Sweet DG, Carnielli V, Greisen G, et al. European Consensus Guidelines on the Management of Respiratory Distress Syndrome - 2016 Update. Neonatology. 2017;111:107–25.

2. Subramaniam P, Ho JJ, Davis PG. Prophylactic nasal continuous positive airway pressure for preventing morbidity and mortality in very preterm infants. Cochrane Database Syst Rev. 2016:CD001243.

3. Doyle LW, Carse E, Adams AM, et al. Ventilation in Extremely Preterm Infants and Respiratory Function at 8 Years. N Engl J Med. 2017;377:329–37.

4. Ciuffini F, Robertson CF, Tingay DG. How best to capture the respiratory consequences of prematurity? Eur Respir Rev. 2018;27 doi: 10.1183/16000617.0108-2017.

5. Chow SSW, Le Marsney, R., Creighton, P., Kander, V., Haslam, R., Lui, K. Report of the Australian and New Zealand Neonatal Network 2015. Sydney ANZNN. 2017.

6. Roehr CC, Schmalisch G, Khakban A, Proquitte H, Wauer RR. Use of continuous positive airway pressure (CPAP) in neonatal units--a survey of current preferences and practice in Germany. Eur J Med Res. 2007;12:139–44.

7. Thebaud B, Goss KN, Laughon M, et al. Bronchopulmonary dysplasia. Nat Rev Dis Primers. 2019;5:78.

8. Dreyfuss D, Saumon G. Ventilator-induced lung injury: lessons from experimental studies. Am J Respir Crit Care Med. 1998;157:294–323.

9. Pereira-Fantini PM, Pang B, Byars SG, et al. Preterm Lung Exhibits Distinct Spatiotemporal Proteome Expression at Initiation of Lung Injury. Am J Respir Cell Mol Biol. 2019;61:631–42.

10. Tingay DG, Pereira-Fantini PM, Oakley R, et al. Gradual Aeration at Birth is More Lung Protective than a Sustained Inflation in Preterm Lambs. Am J Respir Crit Care Med. 2019;200:609–16.

11. Frerichs I, Amato MB, van Kaam AH, et al. Chest electrical impedance tomography examination, data analysis, terminology, clinical use and recommendations: consensus statement of the TRanslational EIT developmeNt stuDy group. Thorax. 2017;72:83–93.

12. Gattinoni L, Caironi P, Pelosi P, Goodman LR. What has computed tomography taught us about the acute respiratory distress syndrome? Am J Respir Crit Care Med. 2001;164:1701–11.

13. Tingay DG, Rajapaksa A, Zonneveld CE, et al. Spatiotemporal Aeration and Lung Injury Patterns Are Influenced by the First Inflation Strategy at Birth. Am J Respir Cell Mol Biol. 2016;54:263–72.

14. Tingay DG, Rajapaksa A, Zannin E, et al. Effectiveness of individualized lung recruitment strategies at birth: an experimental study in preterm lambs. Am J Physiol Lung Cell Mol Physiol. 2017;312:L32–L41.

15. Tingay DG, Polglase GR, Bhatia R, et al. Pressure-limited sustained inflation vs. gradual tidal inflations for resuscitation in preterm lambs. J Appl Physiol. 2015;118:890–7.

16. Sophocleous L, Frerichs I, Miedema M, et al. Clinical performance of a novel textile interface for neonatal chest electrical impedance tomography. Physiol Meas. 2018;39:044004.

17. Armstrong RK, Carlisle HR, Davis PG, Schibler A, Tingay DG. Distribution of tidal ventilation during volume-targeted ventilation is variable and influenced by age in the preterm lung. Intensive Care Med. 2011;37:839–46.

18. Miedema M, van der Burg PS, Beuger S, de Jongh FH, Frerichs I, van Kaam AH. Effect of Nasal Continuous and Biphasic Positive Airway Pressure on Lung Volume in Preterm Infants. J Pediatr. 2013; 164:691-697.

19. Bhatia R, Davis PG, Tingay DG. Regional Volume Characteristics of the Preterm Infant Receiving First Intention Continuous Positive Airway Pressure. J Pediatr. 2017;187:80–8 e2.

20. Tingay DG, Waldmann AD, Frerichs I, Ranganathan S, Adler A. Electrical Impedance Tomography Can Identify Ventilation and Perfusion Defects: A Neonatal Case. Am J Respir Crit Care Med. 2019;199:384–6.

21. Miedema M, Waldmann A, McCall KE, Bohm SH, van Kaam AH, Tingay DG. Individualized Multiplanar Electrical Impedance Tomography in Infants to Optimize Lung Monitoring. Am J Respir Crit Care Med. 2017;195:536–8.

22. Dowse GP, E.; Thomson, J.; Schinckel, N.; Pereira-Fantini, P.; Tingay, D.G. Synchronised inflations generate greater gravity dependent lung ventilation in neonates. J Pediatric. 2020; doi.org/10.1016/j.peds.2020.08.043 [published Online 19 August 2020].

23. Schinckel NF, Hickey L, Perkins EJ, et al. Skin-to-skin care alters regional ventilation in stable neonates. Arch Dis Child Fetal Neonatal Ed. 2020; doi: 10.1136/archdischild-2020-319136 [published Online First: 2020/08/01].

24. Adler A, Arnold JH, Bayford R, et al. GREIT: a unified approach to 2D linear EIT reconstruction of lung images. Physiol Meas. 2009;30:S35.

25. Frerichs I, Dargaville PA, van Genderingen H, Morel DR, Rimensberger PC. Lung volume recruitment after surfactant administration modifies spatial distribution of ventilation. Am J Respir Crit Care Med. 2006;174:772–9.

26. Walsh MC, Yao Q, Gettner P, et al. Impact of a physiologic definition on bronchopulmonary dysplasia rates. Pediatrics. 2004;114:1305–11.

27. Steinberg JM, Schiller HJ, Halter JM, et al. Alveolar instability causes early ventilator-induced lung injury independent of neutrophils. Am J Respir Crit Care Med. 2004;169:57–63.

28. Gattinoni L, Carlesso E, Cadringher P, Valenza F, Vagginelli F, Chiumello D. Physical and biological triggers of ventilator-induced lung injury and its prevention. Eur Respir J Suppl. 2003;47:15s–25s.

29. Tingay DG, Mills JF, Morley CJ, Pellicano A, Dargaville PA. The deflation limb of the pressure-volume relationship in infants during high-frequency ventilation. Am J Respir Crit Care Med. 2006;173:414–20.

30. Miedema M, de Jongh FH, Frerichs I, van Veenendaal MB, van Kaam AH. Changes in lung volume and ventilation during lung recruitment in high-frequency ventilated preterm infants with respiratory distress syndrome. J Pediatr. 2011;159:199–205 e2.

31. Wolf GK, Gomez-Laberge C, Rettig JS, et al. Mechanical ventilation guided by electrical impedance tomography in experimental acute lung injury. Critical Care Medicine. 2013;41:1296–304.

32. Gomez-Laberge C, Rettig JS, Smallwood CD, Boyd TK, Arnold JH, Wolf GK. Interaction of dependent and non-dependent regions of the acutely injured lung during a stepwise recruitment manoeuvre. Physiol Meas. 2013;34:163–77.

33. Zhao Z, Chang MY, Chang MY, et al. Positive end-expiratory pressure titration with electrical impedance tomography and pressure-volume curve in severe acute respiratory distress syndrome. Ann Intensive Care. 2019;9:7.

34. Pham TM, Yuill M, Dakin C, Schibler A. Regional ventilation distribution in the first 6 months of life. Eur Respir J. 2011;37:919–24.

35. Miedema M, de Jongh FH, Frerichs I, van Veenendaal MB, van Kaam AH. The effect of airway pressure and oscillation amplitude on ventilation in pre-term infants. Eur Respir J. 2012;40:479–84.

36. Hough JL, Shearman AD, Jardine L, Caldararo D, Schibler A. Effect of randomization of nasal high flow rate in preterm infants. Pediatr Pulmonol. 2019;54:1410–6.

37. Tingay DG, Lavizzari A, Zonneveld CE, et al. An individualized approach to sustained inflation duration at birth improves outcomes in newborn preterm lambs. Am J Physiol Lung Cell Mol Physiol. 2015;309:L1138–49.

38. Sophocleous L, Waldmann AD, Becher T, et al. Effect of sternal electrode gap and belt rotation on the robustness of pulmonary electrical impedance tomography parameters. Physiol Meas. 2020;41:035003.

39. Schmolzer GM, Bhatia R, Davis PG, Tingay DG. A comparison of different bedside techniques to determine endotracheal tube position in a neonatal piglet model. Pediatr Pulmonology. 2013;48:138–45.

40. Bhatia R, Schmolzer GM, Davis PG, Tingay DG. Electrical impedance tomography can rapidly detect small pneumothoraces in surfactant-depleted piglets. Intensive Care Med. 2012;38:308–15.

