## Supplementary material for "Regional ventilation characteristics during non-invasive respiratory support in preterm infants": Online Supplementary Material

1. **Supplementary Results**

**OSM Table 1.** Role of body position at study on CoV_I-A_.

|  | **Prone (n=31)** | **Supine (n=3)** | **Lateral** | |
| --- | --- | --- | --- | --- |
|  |  |  | **Left side down (n=4)** | **Right side down (n=2)** |
| CoV_I-A_ (%) | 0.6 (-3.5, 10.1) | -2.8 (-2.9, 1.4) | 4.2 (0.1, 17.3) | -8.4, 1.9 |

*All data median (range) or values (right lateral). No statistical comparison was performed due to small group numbers in supine and lateral positions.*

**OSM Table 2.** Relationship between CoV_I-A_ and corrected gestational age, weights and Positive End-Expiratory Pressure (PEEP).

|  | **Slope** | **y intercept** | **R^2^** | **p value** |
| --- | --- | --- | --- | --- |
| **Corrected A** | 0.16 (-0.81, 1.13) | -6.3 (-36.0, 23.4) | 0.003 | 0.74 |
| **Birthweight** | 0.004 (-0.002, 0.010) | -5.3 (-11.5, 0.9) | 0.043 | 0.20 |
| **Weight at study** | -0.002 (-0.007, 0.003) | 1.4 (-5.0, 7.7) | 0.021 | 0.37 |
| **PEEP*** | -0.77 (-2.02, 0.54) | 3.4 (-5.9, 12.7) | 0.046 | 0.24 |

*All data linear regression and error 95% CI.*

** infants only receiving CPAP (n=32)*

**OSM Figure 1.** Study recruitment flow chart.

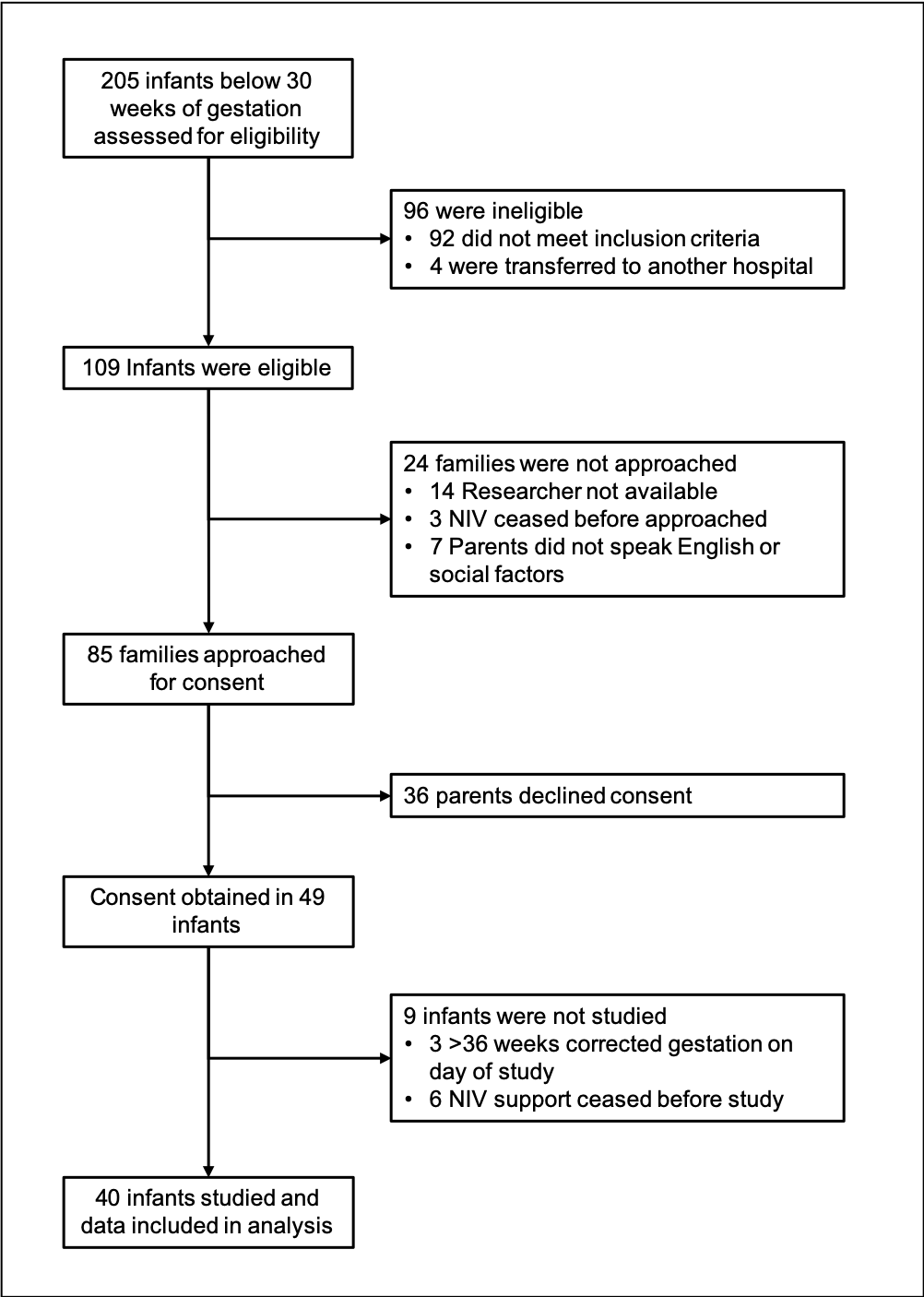

**OSM Figure 2.** Role of body position on location of ideal gravity-dependent centre of ventilation (CoV). To account for the differences in ideal gravity-dependent CoV all CoV data along the non-dependent (0%) to dependent (100%) plane were expressed against the ideal CoV representing uniform ventilation in that position.

| **Position** | **Schematic** | **Non-dependant (0%) to dependant (100%) hemithorax** | **Ideal CoV (non-dependant to dependant)** |
| --- | --- | --- | --- |
| **Supine** | 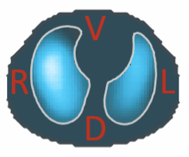 | Ventral lung to Dorsal lung | 55 % |
| **Prone** | 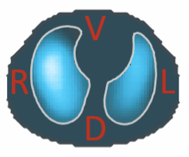 | Dorsal lung to ventral lung | 45 % |
| **Right side down** | 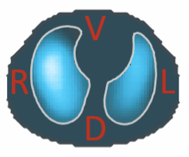 | Left lung to right lung | 54 % |
| **Left side down** | 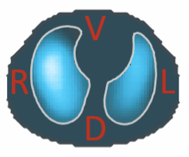 | Right lung to left lung | 46 % |
